# Real-world genetic screening with molecular ancestry supports comprehensive pan-ethnic carrier screening

**DOI:** 10.1101/2022.09.02.22279503

**Authors:** Ryan A. Shewcraft, Mitchell K. Higashi, Yeting Zhang, Jonathan Tyler, Lisa Y. Lau, Bryn D. Webb, Seungwoo Lee, Rajasekar Ramasamudram-Chakravarthi, Teresa A. Cacchione, Alan B. Copperman, Ashley Birch, Marra Francis, Lisong Shi, Lisa Edelmann, Rong Chen, Li Li, Eric Schadt

## Abstract

We characterize the clinical utility and economic benefits of a comprehensive pan-ethnic carrier screening panel that spans 282 monogenic disease conditions in a large, diverse population of 397,540 reproductive health patients. For 142,049 of these patients, we were able to accurately estimate genetic ancestries across 7 major population groups. We examined individual carrier and at-risk carrier couple (ARCC) rates with respect to self-reported and genetic ancestries across ancestry-specific and pan-ethnic panels. Our results show that this comprehensive panel identified >10-times the ARCCs compared with a two-gene pan-ethnic panel and provided a substantial benefit over ancestry-specific screening panels across the major population groups. Finally, we generated a universal cost-of-care model across the monogenic disease conditions represented on the comprehensive pan-ethnic carrier screening panel to demonstrate potential healthcare savings in addition to the demonstrated clinical benefits that could be realized adopting this type of panel as standard of care for all.

## Introduction

Carrier screening is the practice of identifying healthy individuals who may be at risk of having affected offspring with severe monogenic disease and has traditionally been applied to identifying couples whose offspring are at risk for autosomal recessive conditions. However, increasingly genes with dominant and X-linked modes of inheritance are being offered on today’s comprehensive carrier screening panels, which are becoming leveraged more and more as standard of care to maximize reproductive health choices. The increasing utility of comprehensive carrier screening is at least partly driven by an exponential increase in the generation and analysis of genomic data that have led to the identification of hundreds of thousands of DNA variants across thousands of genes that cause severe monogenic disorders. With the improved availability of prenatal diagnostic testing technologies and the increased use of assisted reproductive technologies such as *in vitro fertilization* (IVF) that deploy preimplantation genetic testing for rare monogenic disorders (PGT-M), reproductive health decisions can be more maximally informed.

The acceleration of our understanding of monogenic disorders has been enabled by large-scale genomic studies that seek to associate DNA variation with monogenic disorders, such as the UK Biobank^1–3^, the 1000 Rare Diseases Project^4–7^, large-scale clinical exome sequencing out of the Regeneron Genetics Center^8^, Genomics England^9–12^, and Biobank Japan^13^, as well as improvements in genomics technologies that in 2022 delivered the first complete sequence of the human genome^14^. From these types of large-scale studies, the frequency of individuals with monogenic disorders has been estimated to be in the range of 5-6% across the general population. Further, the characterization of the genetic causes and frequencies of monogenic disorders across diverse population groups have been systematically enhanced by major efforts such as the 1000 Genomes Project, which not only provide global references for human genetic variation across different continental populations^15^, but the combination of a growing number of modern and ancient genomes have helped lead the way to a single, unified genealogy defining the ancestral relationships among all living and past humans^16^. With this increasing characterization of monogenic disorders and rare disorders more generally, comes more accurate estimates of the significant direct and indirect costs, where in the US alone costs are estimated to be approaching $1 trillion annually for rare disorders^17^, comprised roughly of $449 billion in direct medical costs^17^,with a significant portion of these costs attributable to genetic diseases that manifest in the NICU and pediatric settings^18^. Further, parents of affected neonates and children typically face a prolonged diagnostic odyssey^19–21^ in addition to significant caregiver burden^22^ and financial costs^17^.

The use of more comprehensive preconception carrier screening has become one way in which reproductive health couples can benefit from the increased understanding of monogenic disorders coming out of the genomics revolution, providing for enhanced reproductive decision making that can dramatically reduce the probably of conceiving a child with a severe monogenic disorder. Practice guidelines have also begun to acknowledge this increased understanding of monogenic disorders and their frequency throughout different population groups. For example, this past year substantially revised practice guidelines for carrier screening were released by American College of Medical Genetics (ACMG)^23^, with strong support now expressed for the expansion of carrier screening across more than 100 autosomal recessive and X-linked conditions, with an emphasis on providing carrier screening options in an ethnic and population neutral way to promote greater inclusivity and diversity with respect to access to this important emerging standard of care. The pan-ethnic positioning of these guidelines has been further supported by the introduction of molecular ancestry, with one recent study of genetic ancestry carried out on 93,410 individuals who had received carrier screening, discovering that 9% of the individuals analyzed have >50% genetic ancestry from lineages that are inconsistent with their self-report ancestry^24^.

The medical policies of some health insurers and many physician practices cite unproven medical necessity and insufficient evidence of efficacy for comprehensive carrier screening testing as among some of the reasons to not promote and cover the cost of its use^25–27^. Further, medical policies among the major national health insurers are broadly inconsistent with respect to qualifying as medically necessary ethnicity-specific carrier screening panels as medically necessary, approving for some well-studied population groups such as the Ashkenazi Jewish (AJ) population, while not approving such screening for other population groups where the risks are comparable to that of the AJ population. In the diagnostic screening context, clinical utility is established by demonstrating reduced adverse health outcomes by adopting interventions informed by the testing results^28^. More specifically, for carrier screening, clinical utility can be established by demonstrating an impact on reproductive decision making as a result of positive test results ^29^. Several large-scale studies have demonstrated impacts on reproductive decision making through the use of comprehensive carrier screening, where in cases a positive, at-risk carrier couple has been identified prior to establishing a pregnancy, decisions were subsequently made to establish a pregnancy using IVF combined with preimplantation genetic testing or to forgo pregnancy and adopt a child. In cases where a pregnancy had already been established, positive expanded carrier screening can lead to prenatal testing, facilitating decision making around continuation of the pregnancy^30–33^. However, in these studies the impact on decision making was established via surveys post pregnancy, as opposed to direct assessment of outcomes through the patient medical records. One recent study, albeit smaller than the above studies, more directly established clinical validity and utility of comprehensive carrier screening by following 15 carrier couples, out of several hundred who had been tested^34^. In all cases the couples modified their reproductive planning as a result of the carrier screening results, to include decisions to achieve pregnancy through IVF, resulting in the generation of euploid/unaffected embryos that in some cases were successfully transferred and resulted in the birth of unaffected babies.

To provide further direct support of the clinical validity, utility, and value of comprehensive carrier screening, here we examined the results of 397,540 patients who received a comprehensive panethnic carrier screening test (283 gene, CS-283) as part of their reproductive health journey, characterizing interpretation of the results with respect to carrier frequency rates, carrier couple rates, and for a subset of carrier couples, an assessment of the direct impact on reproductive health decisions and pregnancy outcomes. Importantly, for 142,049 of the patients, we inferred genetic ancestry using low-pass whole genome sequencing data to compare genetically inferred ancestries with self-reported ancestries, and to characterize the impact of ancestry-specific carrier screening panels with pan-ethnic carrier screening panels. We compare the CS-283 detection rates and subsequent impact on reproductive decision making with a two-gene standard pan-ethnic panel (SPEP) supported by current ACOG guidelines and all major national payers, as well as an ACMG-guideline-informed panel and ancestry-specific panels across 7 major population groups, to further highlight the utility of a pan-ethnic approach to comprehensive carrier screening. We identified > 60% of those tested with the CS-283 panel as carriers of pathogenic variants in at least one of the genes represented on this panel, versus 3% that would have been identified as carriers using the SPEP. Further, we show that 3.9% of those couples screened were determined to be at-risk carrier couples (ARCC) at very high risk (25% or greater) of passing on one of the 283 rare monogenic disorders represented on the CS-283 panel to their offspring, versus a 0.3% ARCC detection rate that would have resulted using SPEP (a 13-fold reduction compared to the ARCC rate achieved with CS-283). We further show that genetic ancestry inferred from low-pass whole genome sequencing resulted in a more robust characterization of genetic risk profiles compared to self-reported ancestry. Based on the genetic ancestry profile for each individual, detection of the individual carriers or at-risk carrier couples from CS-283 are substantially increased compared to ancestry-based gene panels, demonstrating a significantly increased benefit for couples through expanded reproductive health decision-making choices. Importantly, we demonstrate this clinical benefit explicitly in the REI setting, where the identification of at-risk carrier couples in one large Reproductive Endocrinology and Infertility (REI) center, led to the identification of embryos with the corresponding monogenic disease genotypes informed by the CS-283 screening, that were consequently not considered for implantation. We estimate the potential for significantly decreased healthcare costs resulting from increased disease avoidance utilizing pan-ethnic comprehensive carrier screening advocated by the ACMG, compared to less comprehensive pan-ethnic screening tools such as SPEP currently indicated as the standard of care by ACOG and many major national health insurers.

## Results

### Comprehensive carrier screening on a large patient population

We assembled a set of 397,540 patients who received a comprehensive carrier screening test that covered 283 genes (CS-283) and spanned 282 monogenic disease conditions as part of the standard of care they received along their reproductive health journeys (Figure 1A). From these data we sought to characterize individual patient carrier rates, at-risk carrier couple rates, the impact of genetic ancestry on the resulting genetic risk profiles, and the impact on reproductive decision making and pregnancy outcomes (Figure 1B). We carried out these characterizations with respect to the full 283-gene panel, a 2-gene subset of this panel representing a standard carrier screening pan-ethnic panel (SPEP), supported by current ACOG guidelines for all patients who are pregnant or thinking of becoming pregnant, and a 77-gene ACMG-guidelines-based panel covering genes in addition to those in SPEP associated with severe monogenic diseases (ACMG recommended a panel of 113 genes for pan-ethnic carrier screening to be applied as the standard of care for all reproductive health age women, and of those 113 genes, 77 were overlapping with the CS-283 panel)^23^. SPEP covers two of the most frequent and severe monogenic disorders: Cystic Fibrosis (*CFTR)* and Spinal Muscular Atrophy (*SMN1)*, where both of these genes are recommended by ACOG guidelines^35^. The 397,540 patients tested spanned a broad reproductive age range (Figure 1C) and self-reported ethnicities (Figure 1D). Of those, 59% were female, consistent with clinical practice where the female partner often receives carrier screening first and then the male partner is sought for testing if a positive test results. Three-quarters of the patients tested (306,226; 77%) had their carrier screening tests ordered through Reproductive Endocrinology and Fertility (REI) clinics, while the remaining patients (91,314; 23%) had tests ordered through Maternal Fetal Medicine (MFM) or general Obstetrician-Gynecologist (Ob-Gyn) practices (Figure 1A, 1E). Given IVF accounts for only 2% of all infants born in the U.S.^36^, the extreme bias in our CS-283 test population for patients undergoing IVF, a highly medicalized reproductive process with frequent laboratory and imaging testing before and after egg retrieval and embryo implantation, reflects the more rapid uptake of advanced comprehensive carrier screening as the standard of care in this community.

**Figure 1:**
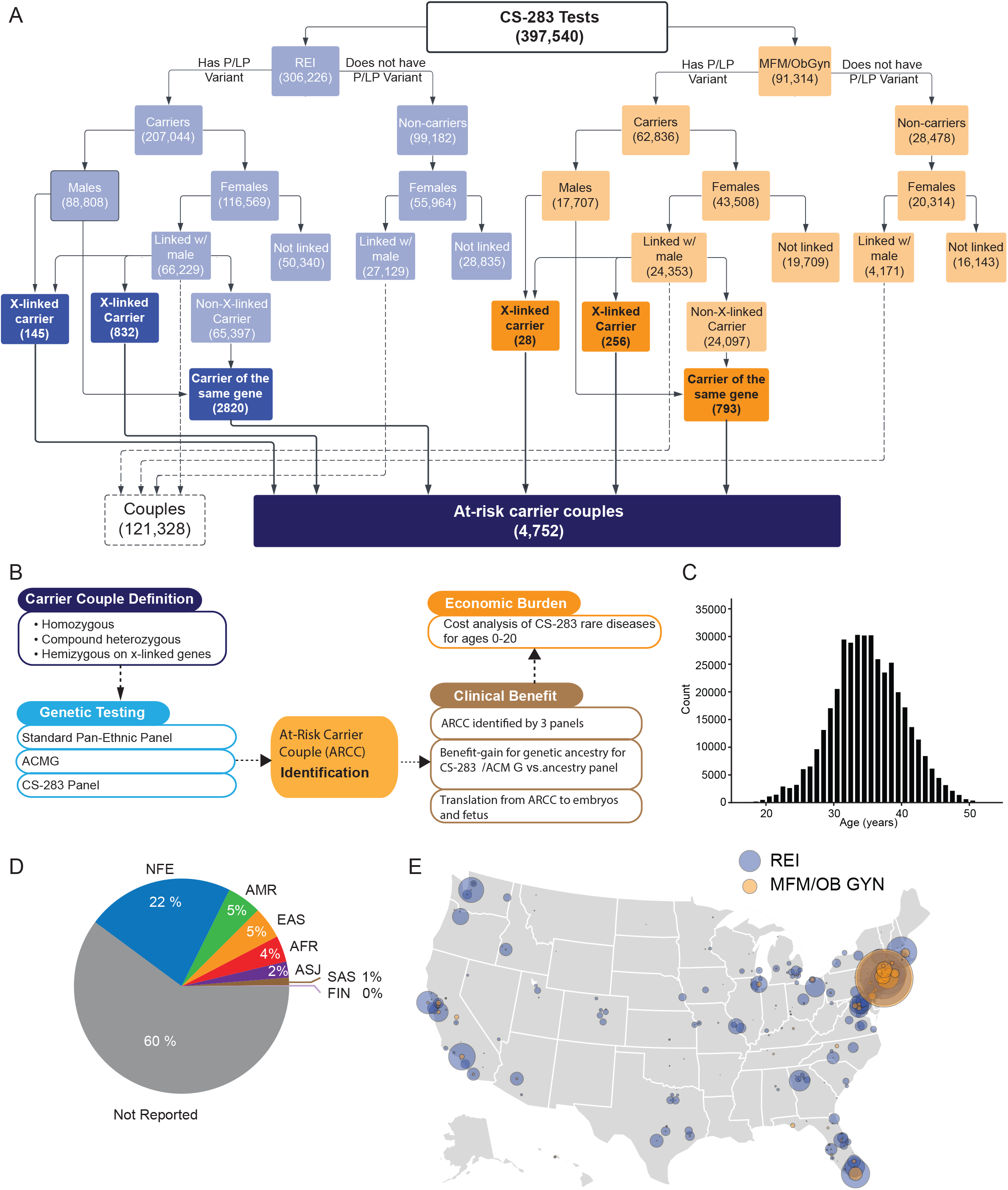
Cohort demographics. **(A)** Flowchart for determining carrier status for both REI and MFM/Ob-Gyn practices. **(B)** Outline of logic and workflow. The number of individual carriers and carrier couples were identified by several carrier screening panels was compared to estimate both the clinical benefit of pan-ethnic carrier screening and the economic burden of diseases covered in the larger panel. **(C)** Age distribution at the time carrier screening was ordered for individuals who received CS-283 screening. **(D)** Distribution of self-reported ancestries mapped to lpWGS-based ancestry categories for individuals who received CS-283 screening: NFE: Non-Finnish European, ASJ: Ashkenazi Jewish, AMR: Admixed American, EAS: East Asian, AFR: African, SAS: South Asian, FIN: Finnish. **(E)** Geographic distribution of locations of practices where carrier screening was ordered split by (purple) and MFM/Ob-Gyn (yellow). The size of the circle corresponds with the number of individuals. Locations were grouped by and presented at the county level.

The geographic distribution of carrier screening tests that were ordered by REI centers or MFM/Ob-Gyn clinics is depicted in Figure 1E. We grouped practices at the county level and then plotted the total number of tests ordered in each county by practice type. In general, we found that a substantial number of tests were ordered around major cities, particularly for REI practices. While most of the tests were ordered in coastal states, patients represented in our test population spanned 47 of the 50 U.S. states (South Dakota, Louisiana, and Alaska were the only states not represented). In the New York City tristate area and Florida, the proportion of patients coming from REI and MFM/Ob-Gyn practices were similar, while at most other locations most orders were from REI clinics. Overall, the US population is broadly represented in our comprehensive carrier screening test population.

### Impact of ancestry determination on genetic risk prediction in test population

The test population is ethnically diverse, representing all major population groups across the United States (Figures 1D & E). Given the self-reported nature of ancestry provided on the physician test order forms, it is noteworthy that a substantial proportion of the test population (60%) did not report their ancestry (Not reporting; NR). An NR designation results if the patient does not provide ancestry information to the ordering physician, the patient is unsure of their ancestry, or the ordering physician simply omits this information. Thus, despite the importance of ancestry information for assessing carrier risk given widespread differences in carrier rates and disease prevalence among different population groups, this information is typically missing on the patient at the time the carrier screening test is ordered. For the remaining patients, self-reported ancestry was mapped to one of the seven population groups described below.

Complementing self-reported ancestry is genetic ancestry, which can be inferred from the type of genome-wide DNA variation information imputed from low-pass whole genome sequencing (lpWGS) data. Genetic ancestry provides a data-driven way to accurately estimate the composition of individual patients with respect to different population groups, whereas self-reported ancestry may be confounded by sociocultural factors. In the absence of comprehensive pan-ethnic carrier screening for all, an accurate estimate of genetic ancestry is critical for ensuring monogenic disorders overrepresented in a patient’s ancestral population groups, are represented in the carrier screening test administered to the patient. For example, the population prevalence of Gaucher disease is nearly 100-times higher in the Ashkenazi Jewish (ASJ) population compared to all other continental population groups^37^, and thus patients reporting ASJ ancestry should be tested with a carrier screening panel such as ACMG that includes the *GBA* gene that causes Gaucher disease, as opposed to SPEP, which does not include the *GBA* gene. Accurate ancestry information is also necessary for ensuring the most reliable residual risk calculations for the patient.

For 142,049 of the 397,540 (35.7%) patients in the test population, lpWGS data were generated as an integrated component of the carrier screening test administered to infer genetic ancestry for more accurate residual risk calculations. The lpWGS data were used to model each individual as a mixture of 7 different major population groups: Finnish (FIN), Ashkenazi Jewish (ASJ), African (AFR), South Asian (SAS), East Asian (EAS), admixed American (AMR), and non-Finnish European (NFE). With reference genomes for each group, an admixture proportion inference approach was employed to estimate the percentage of the genome in each individual that was inherited from each population group (See Methods). Whereas self-reported ancestry is categorical and often unidimensional, genetic ancestry exists on a spectrum across all population groups. Therefore, we would expect varying degrees of discordance between genetic and self-reported ancestries that may impact genetic risk scores.

To explore this, we examined genetic risk for 6 of the 7 population groups (ASJ, AFR, EAS, SAS, AMR, and NFE; FIN was excluded due to small sample size) to obtain reliable estimates. For each population group, we used 25% as an appropriate threshold to classify whether each patient with lpWGS data in the test set had a significant percentage of their genome derived from the indicated population group or not (Figure 2; See Methods). A 25% ancestry threshold is consistent with inheriting DNA from a first or second degree relative descended from the indicated population group, a common criterion applied by physicians, genetic counselors, and health insurers for considering whether ancestry-specific screening is warranted. The distributions of genetic and self-reported ancestry proportions for each population group across all individuals with a non-zero ancestry proportion estimated from the lpWGS data and whose self-reported ancestry was not indicated as “Not reported”, are given in column (i) of Figure 2. The self-reported ancestry proportions for those identified as having genetic ancestry for a given population group are provided in column (ii) of Figure 2.

**Figure 2:**
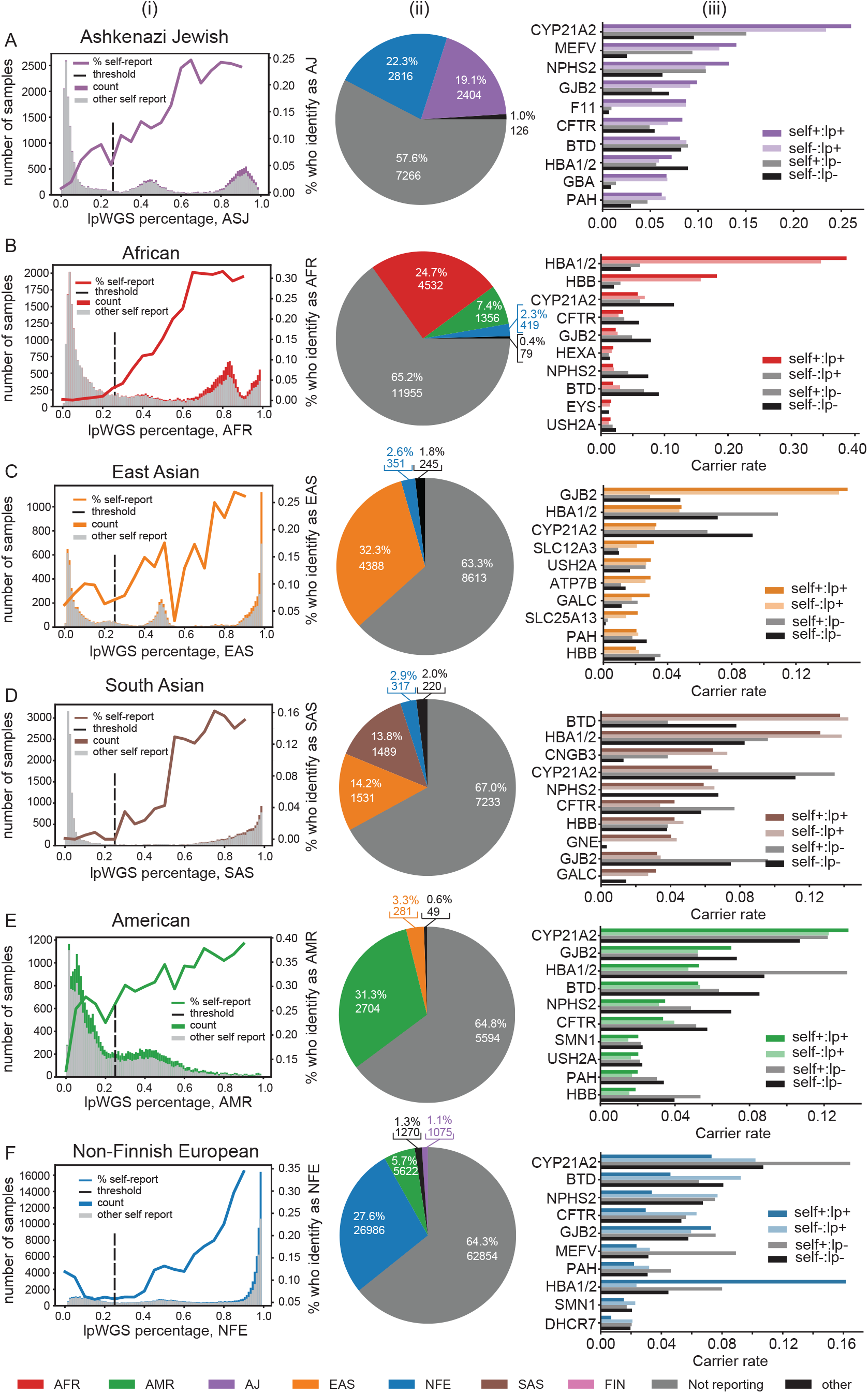
Self-reported ancestry in comparison to lpWGS-based ancestry. **(Ai)** Distribution of lpWGS-ancestry proportions for all individuals with non-zero ancestry for the ASJ ancestry category. The purple area in the histogram shows the number of patients with ASJ self-reported ethnicity. The gray area in the histogram shows the number of patients with self-reported ethnicity other than ASJ. The solid line shows the overall percentage of individuals in each lpWGS proportion bin who self-reported as ASJ. The dotted line shows the binary threshold determined as 25% cutoff. **(Aii)** Breakdown of self-reported ethnicities for individuals whose lpWGS proportion was greater than the ASJ threshold. The percentage of each ethnicity when not including “not reporting” is shown in parentheses. **(Aiii)** Individual carrier rates for most frequent genes split by self-reported ethnicity/ancestry status. Self+:lp+: individuals who have both lpWGS-based ancestry proportion above the ASJ threshold and also self-reported ethnicity as ASJ; Self-:lp+: individuals whose lpWGS-based ancestry proportion was above the ASJ threshold but self-reported ethnicity was not; “Self+:lp-”: individuals whose self-reported ethnicity was ASJ but lpWGS-based ancestry proportion was below the ASJ threshold; “Self-:lp-”: individuals in which self-reported ethnicity was not ASJ and their ancestry proportion is below the ASJ threshold. **(B)** Same as (A) for AFR. **(C)** Same as (A) for EAS. **(D)** Same as (A) for SAS. **(E)** Same as (A) for AMR. **(F)** Same as (A) for NFE.

From column (i) in Figure 2, we observe that the self-reported ancestry is more likely to match the genetic ancestry as the proportion of genetic ancestry increases. However, we observed substantial differences. For example, we identified 12,612 patients with ASJ ancestry (25% vertical dashed threshold line in Figure 2A(i)). Strikingly, of these patients with ASJ genetic-based ancestry, 57.6% did not self-report their ancestry, followed by 22.3% who self-reported with NFE ancestry instead of ASJ ancestry. The 19.1% who self-reported with ASJ ancestry and who were inferred as having ASJ genetic ancestry, comprised only 45.0% of those patients with genetic ASJ ancestry who also self-reported their ancestry (Figure 2Aii). Thus, more than three-quarters of the test population with substantial ASJ heritage, were not indicated in the physician’s order for the carrier screening test as having ASJ ancestry (Figure 2Aii). A similar pattern was observed for those classified with genetic SAS ancestry (Figure 2Di & ii), with only 13.8% of those with genetic SAS ancestry, self-reporting with SAS ancestry, which represents only 41.9% of those with a self-reported ancestry. Sixty-seven percent of those classified with genetic SAS ancestry, had not self-reported in the test order form. For those classified with genetic AFR and EAS ancestries (Figures 2Bi & ii and 2Ci & ii), when provided, the self-reported ancestry matched the genetic ancestry 71.0% and 88.0% of the time, respectively, although both groups still had high NR rates for self-reported ancestry (65.2% and 63.3%, respectively). Similar results hold for the AMR and NFE population groups (Figures 2E and 2F, respectively).

To illustrate the differences in genetic risk predictions between genetic and self-reported ancestries, we examined the top 10 genes from the CS-283 panel ranked by individual carrier rates in each of the 4 population groups (column (iii) in Figure 2). The 10 top-ranked genes for each population group were all contained within the set of genes known to be significantly overrepresented in the population group (Supplementary Table 9). To characterize the concordance/discordance between self-reported and genetic-ancestry and the impact on carrier rates, we estimated the proportion of patients for each population group across the 4 possible combinations of self-reporting (self+ or self-) and genetic (lp+ or lp-) ancestry calls. The empirical carrier rates for all 4 possible combinations of self-reported:genetic ancestry (self+/-:lp+/-) results with respect to the indicated population group across the top 10 genes, are plotted in column (iii) of figure 2. In nearly all cases, the carrier rates for the genetic ancestry regardless of the self-reported ancestry, best reflected the expected overrepresentation of carrier rates with respect to the indicated population group, compared to the self-reported ancestry. For example, for those classified as EAS, *GJB2* is the top ranked gene known to be overrepresented in the EAS population^37^, and the patients classified with genetic EAS ancestry well captured this high carrier rate in the test population, even in those individuals who did not self-report as EAS. Further, those patients not classified as having genetic EAS ancestry had substantially lower carrier rates, as expected (column (iii), Figure 2C).

This type of pattern held up across each of the population groups. For example, in the ASJ population, genetic ancestry more accurately identified genes known to have high carrier rates in the ASJ population that otherwise would have been missed in those patients not self-reporting as having ASJ ancestry (note the *self-:lp+* and *self+:lp+* bars in column (iii), Figure 2A). In fact, the correlation between the individual carrier rates for the top 10 genes in the ASJ population listed in Figure 2A between individuals who self-report as ASJ and those who did not self-report as ASJ, when these individuals were classified as having genetic ASJ ancestry, was 0.989 (p=5.64×10^−8^), whereas the correlation between individuals with genetic ASJ ancestry and those without genetic ASJ ancestry when these individuals self-reported as having ASJ ancestry was 0.83 (p = 0.003). We classified 18,341 patients (12.9% of those tested with lpWGS data) as having genetic African ancestry (column (i), Figure 2B), and then similarly investigated the top 10 genes ranked by individual carrier rates in this population group (column (iii), Figure 2B). Among the top ten genes, the *HBB* and *HBA1/HBA2* genes are known to be high-prevalence risk genes in the African population group (Suppleementary Table 9). Again, the correlation between the individual carrier rates between the (*self-:lp+)* and *(self+:lp+)* groups was high (r=0.998, p=1.61×10^−9^), compared to the correlation between the (*self+:lp+*) and (*self+:lp-*) groups (r=0.22, p = 0.57), demonstrating the robustness of utilizing lpWGS to determine ancestry.

For the EAS population group, 13,597 (9.6%) individuals were classified as having genetic EAS ancestry (column (i), Figure 2C). Patients with genetic EAS ancestry had the highest concordance with self-reported ancestry with 88.0% of those patients with genetic EAS ancestry who self-reported an ancestry, self-reporting as EAS (column (ii), Figure 2C). The top 10 genes ranked by individual carrier rates are shown in column (iii) of Figure 2C, and the correlation between the (*self-:lp+*) and (*self+:lp+*) groups is 0.995 (p=3.15×10^−9^), compared to a correlation of 0.14 (p = 0.71) between the (*self+:lp+*) and (*self+:lp-*) groups. As with the ASJ and AFR groups, the top 10 ranked genes for EAS were among the genes most enriched in the EAS population group (Supplementary Table 9). Finally, 10,790 patients were classified as having genetic SAS ancestry (column (i), Figure 2D). Individuals with genetic SAS ancestry self-reported with EAS ancestry (14.2%) slightly more often than they self-reported with SAS ancestry (13.8%; column (ii), Figure 2D). This result is perhaps not surprising given the demographic and shared cultural background of the EAS and SAS population groups. For SAS, the correlation between the (*self-:lp+*) and (*self+:lp+*) groups was 0.99 (p=1.9×10^−6^), compared to a correlation of −0.14 (p = 0.77) between the (*self+:lp+*) and (*self+:lp-*) groups. As with the other population groups, the top 10 genes empirically ranked by individual carrier rates in the test population (column (iii), Figure 2D), were among the genes most enriched in the SAS population group (Supplementary Table 9).

### Individual carrier detection rates across different carrier screening panels and ancestries

To identify individual carriers, we examined the CS-283 screening results in the test population, where a patient was identified as a carrier if they were observed with a heterozygous pathogenic/likely pathogenic variant in an autosomal recessive gene or with a hemizygous variant in an X-linked gene, for at least one of the 283 genes represented in the CS-283 panel. Overall, we identified 269,880 carriers out of 397,540 patients tested (67.8%). Individuals with ASJ ancestry had the highest carrier rate (82%), followed by those with NFE ancestry (71%). Nevertheless, for each population group the carrier rate was > 60% on the CS-283 panel.

To compare the carrier rates observed with respect to the CS-283 panel to the rates observed from the SPEP and ACMG panels, we recomputed the carrier rates by restricting analyses to the genes on the SPEP and ACMG panels, and then classified each individual identified as a CS-283 carrier as 1) an SPEP carrier if the carrier gene was on the SPEP panel, 2) an ACMG-specific carrier if the carrier gene was on the ACMG panel but not SPEP, and 3) a CS-283-specific carrier if the carrier gene was in the CS-283 but not the SPEP or ACMG panels. Of all carriers detected, 3% were SPEP carriers (carriers with respect to the *CFTR* or *SMN1* genes), 42% were ACMG-specific carriers, and 55% were CS-283-specific carriers (Figure 3A), so that the majority of carriers identified were identified only as a result of having a more comprehensive carrier screening test. The genes with the highest carrier rates across the test population are shown in Figure 3B, and a full list of the individual carrier rates can be found in Supplementary Table 5.

**Figure 3:**
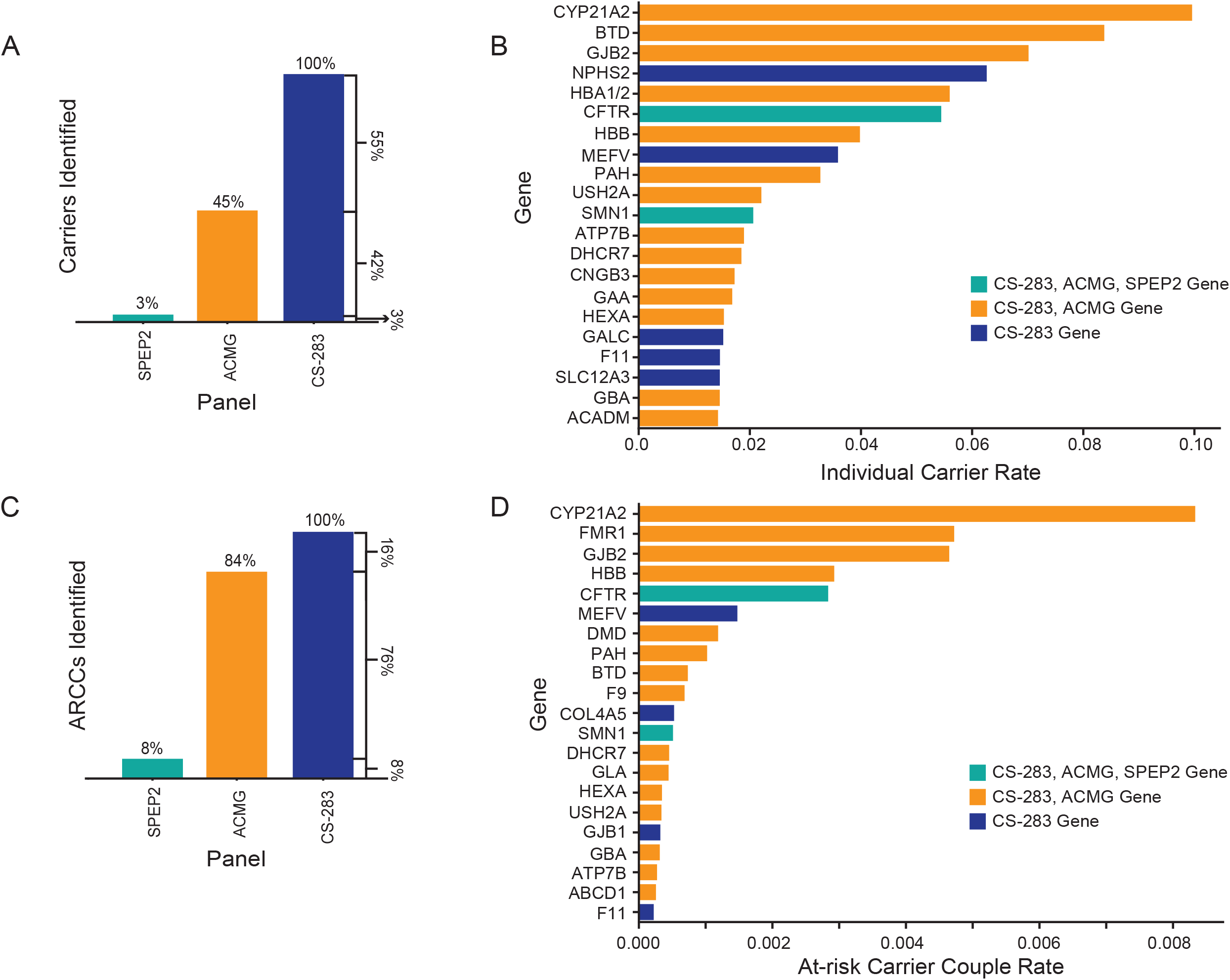
Individual carriers and at-risk carrier couples. **(A)** Breakdown of carriers identified in the test population whose carrier status could be determined by the SPEP, ACMG, or CS-283 panels. For the proportion reported for CS-283, this represents carriers for genes on the CS-283 panel that were not in the SPEP or ACMG panel. **(B)** Individual carrier rate split by gene. The color corresponds to the first panel in which the gene is present, e.g., teal color represents genes in the SPEP, ACMG, and CS-283 panels. **(C)** Breakdown of at-risk carrier couples identified in the test population whose carrier status could be determined by the SPEP, ACMG, or CS-283 panels. **(D)** The at-risk carrier couple rate split by gene. The color corresponds to the first panel in which the gene is present, i.e., teal bars represent genes in all three panels, orange in ACMG and CS-283, and blue, genes only in CS-283.

Given the scaled genetic ancestry results for a significant proportion of the test population, we were able to directly assess the impact of the ACMG and CS-283 panethnic panels on carrier rates as a function of ancestry-specific panels representing the most frequent monogenic disorders for each population group. Given ACOG guidelines and the medical policies of many major health insurers consider more comprehensive carrier screening for some population groups such as ASJ as standard of care, but not for others. In contrast, ACMG guidelines advocate for comprehensive panethnic carrier screening for all over ancestry-specific carrier screening. Thus, we wanted to examine what standard of care positions are best supported by the data.

Towards this end, we constructed ancestry-specific panels for each of the 7 major population groups by identifying those genes with carrier frequencies of pathogenic or likely pathogenic variants ≥ 1/100 for monogenic disorders in each population group (Supplementary Table 9), across the 142,049 patients in the test population with lpWGS data. For each of these panels we then calculated a normalized ratio of CS-283-specific carrier rates over the ancestry-specific carrier rate for each population groups (see Methods for additional details). The sign of this ratio indicates if the CS-283-specific panel or ancestry-specific panel provided greater benefit (positive or negative ratio, respectively). The magnitude of the ratio is an indicator of the degree of benefit provided by the more informative panel. For comparison to the pan-ethnic CS-283 panel, we also calculated this same ratio with respect to the ACMG-informed pan-ethnic panel (77 genes). Across all population groups, the normalized ratios were all positive, ranging from 0.14-1.66 for the CS-283 panel over ancestry-specific panels (blue bars, Figure 4A), and −0.08 to 0.85 for the ACMG panel (orange bars, Figure 4A). For example, the ratio for the ASJ population group was 0.14, indicating that an additional 14% of patients relative to those identified as carriers from the ASJ-specific panel, were identified as carriers for pathogenic variants in genes that were not represented on the ASJ-specific panel. For non-ASJ ancestries, the benefits of the pan-ethnic CS-283 panel are more striking. For example, for the AMR population group the ratios are 1.66 (0.85) for the CS-283 (ACMG) panels, indicating that an additional 166% (85%) of patients relative to those identified as carriers using the AMR-specific panel, were identified as carriers of pathogenic variants in genes not covered by the AMR-specific panel (a > 10-fold higher benefit achieved through the CS-283 for the AMR population group compared to the better characterized ASJ population group). The benefit achieved across all non-ASJ population groups was greater more than 3-fold higher compared with the ASJ population group, suggesting these other population groups are under studied relative to the ASJ population group, and thus achieve a comparable standard of care to the ASJ group only through a panethnic carrier screening panel.

**Figure 4:**
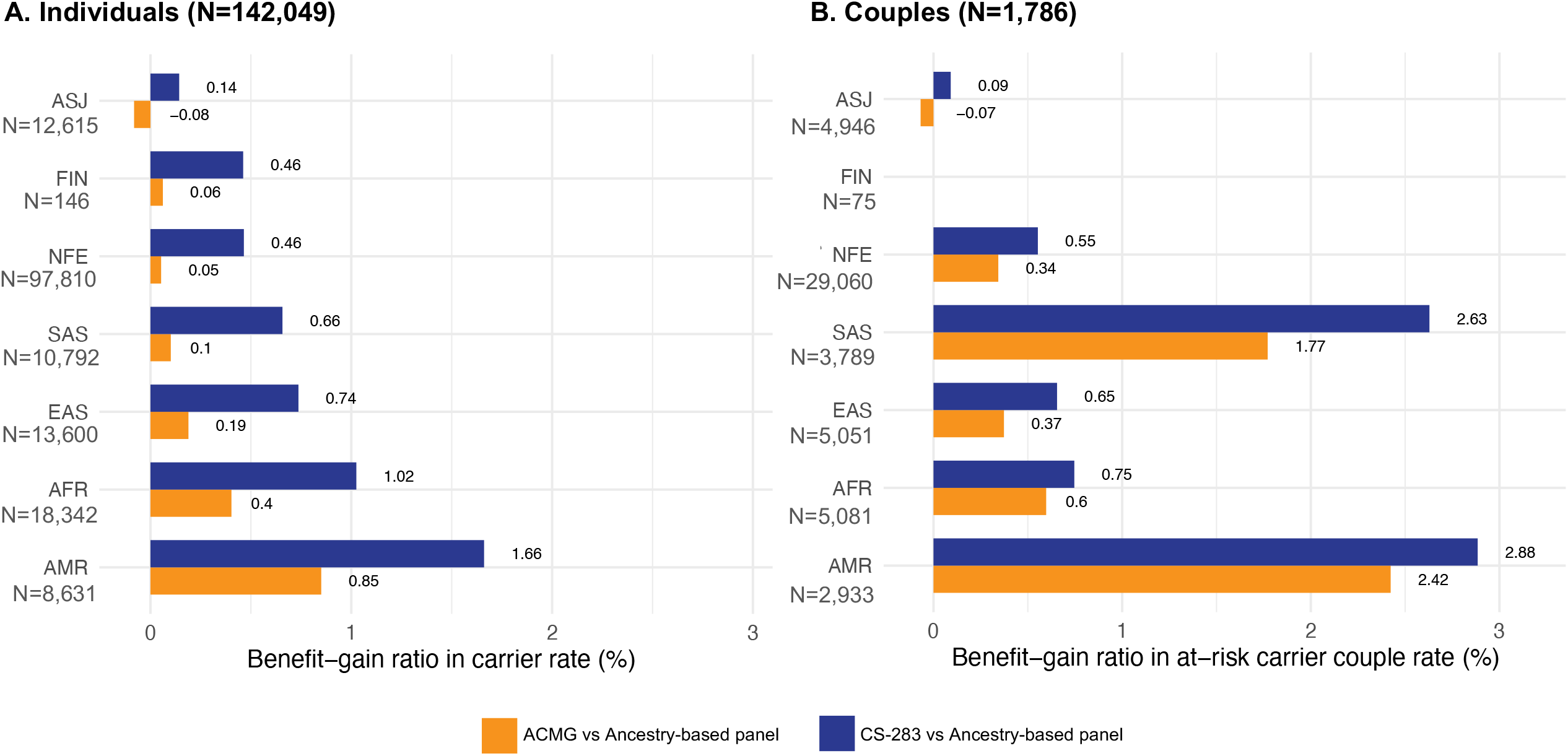
Benefit-Gain ratio in carrier rate when using the CS-283 panel and ACMG-informed panels relative to ancestry-based panels. **(A)** Individual carriers **(B)** At-risk carrier couples

### At-risk carrier couple detection rates across different carrier screening panels and ancestries

For the test population, female and male reproductive partners were able to be connected a significant proportion of the time, and thus at-risk couple carrier (ARCC) rates could be empirically estimated based on real-world coupling data. From the 397,540 patients in the test population, we had information definitively linking partners for 243,764 of them (62.0%), resulting in the identification of 121,328 reproductive health couples. Of these couples, 94,794 (78.1%) received testing through REI clinics, consistent with the approximate overall 3:1 ratio of patients in the test population receiving testing from REI practices relative to those receiving testing in MFM/Ob-Gyn practices. We used these data to empirically estimate ARCC rates (Figure 1B). In total we identified 4,752 ARCC (3.9%) (Figure 1B).

With respect to the SPEP, ACMG-specific, and CS-283-specific panels, 8% of the 4,752 ARCCs were identified from the SPEP, an additional 76% were identified from the ACMG panel, and the remaining 16% were captured only because of the more comprehensive nature of the CS-283 panel (Figure 3C). The CS-283 and ACMG pan-ethnic panels identified 11.5-times and 10.5-times as many ARCCs, respectively, compared to the two-gene SPEP panel (Table 1, Supplementary Table 6).

From the ARCCs identified from the CS-283 panel, we summarized the most common genes represented (Figure 3D). The most frequently affected gene was *CYP21A2*, which was the most frequently affected gene in individual carriers as well. We have included couples that have classic CAH and non-classic CAH in the ARCC. The non-classic CAH causing genetic variants are listed in Supplementary Table S3. We note that NC-CAH individuals develop symptomatic hyperandrogenism that requires longer-acting glucocorticoids treatment in adolescents ^38^. A full list of the ARCC rates can be found in Supplementary Table 7.

As with the individual carrier rates, we leveraged the genetic ancestry information to calculate a normalized ratio to assess the benefit of a pan-ethnic carrier screening panel CS-283 over ancestry-specific panels (blue bars, Figure 4B) and the ACMG-informed panel (orange bars, Figure 4B). Of the 4,752 ARCCs, 1,786 had at least one partner with lpWGS data. Except for the FIN population group (given the small sample size of this population group in the test population, there were no FIN ARCCs identified by a FIN-specific panel), the normalized ratios for the CS-283-specific (ACMG-specific) panels ranged from 0.09 (−0.07) for the ASJ population group to 2.88 (2.42) for the AMR population group. The benefit for ARCC detection with the CS-283 and ACMG panels in non-ASJ population groups was more than 6- and 4-times higher, respectively, compared to the ASJ population group, with the AMR group having a CS-283-specific detection benefit that was 32-times higher than that achieved by the ASJ population group, again supporting that these non-ASJ ancestries are under studied and only through pan-ethnic carrier screening can they achieve the same benefit as the ASJ population group.

### Pan-ethnic screening with the CS-283 panel impacts embryo implantation and pregnancy outcomes

We retrieved the medical record data for 117 ARCCs identified at a single, large REI clinic (out of the total 4,752 ARCCs we identified across all REI, MFM, and Ob-Gyn clinics in our study), and identified 43 who underwent preimplantation genetic testing (PGT-M) as a consequence of having been designated as an at-risk carrier couple. An embryo was considered unaffected if the PGT results indicated it as a noncarrier, carrier, or was a “no call”. Each couple produced from 1 to 40 embryos, and 72% (31 out of 43) had at least one affected embryo defined by PGT, where the PGT was designed based on the CS-283 carrier results. Among the 31 ARCCs with affected embryos, 10.5% involved one of the two genes in SPEP, 58% (18 out of 31) involved one of 11 genes captured by the ACMG gene panel but not SPEP, whereas 32% (10 out of 31) involved one of 6 genes captured by the CS-283 panel but not SPEP or ACMG (Table 2). These detection rates for the pan-ethnic CS-283 and ACMG panels are 6-times and 3.3-times higher, respectively, than the SPEP (Table 2). The affected embryos were not considered further for implantation and were discarded. Among the other 74 ARCCs, it is of note that 27 (37%) declined PGT-M due to being at risk for less severe monogenic conditions: non-classic CAH (N=20), non-syndromic hearing loss (*GJB2*-related) (N=4), and familial Mediterranean fever (N=3). These results reflect the enhanced decision making provided by the CS-283 panel, where in cases of risk of less severe diseases, couples may opt for medical treatment of the condition in affected offspring with shorter time to diagnosis and matching to the most appropriate treatment.

**Table 1:**
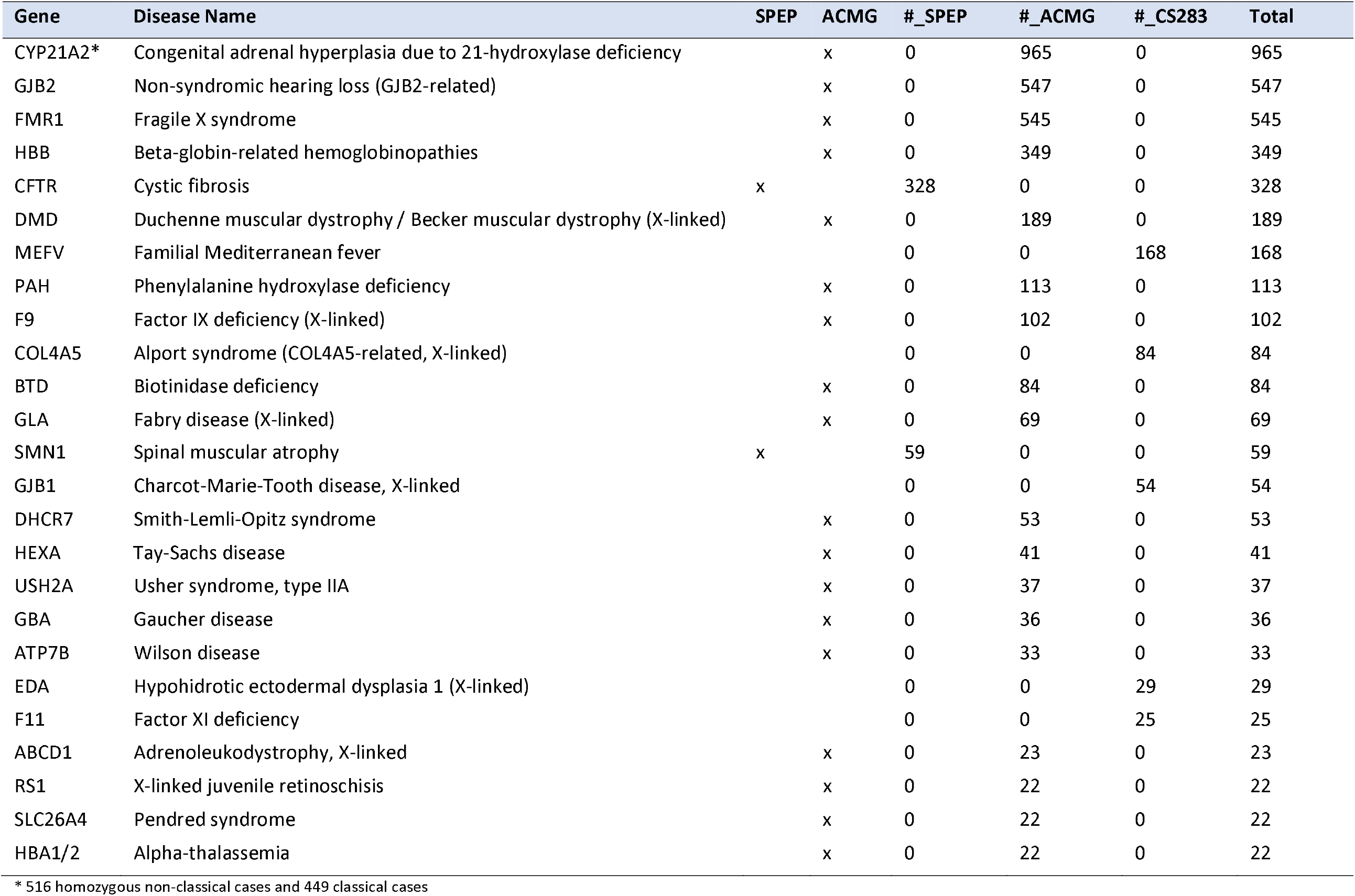
At-risk carrier couple (ARCC) counts (top 25) by test panels, including SPEP, ACMG genes, and CS-283

**Table 2:** Embryos with affected diseases from at-risk carrier couples

| Gene | Disease Name | SPEP | ACMG | #_SPEP | #_ACMG | #_CS283 | Total |
| --- | --- | --- | --- | --- | --- | --- | --- |
| GJB2 | Non-syndromic hearing loss (GJB2-related) |  | x |  | 6 |  | 6 |
| MEFV | Familial Mediterranean fever |  |  |  |  | 4 | 4 |
| PAH | Phenylalanine hydroxylase deficiency |  | x |  | 3 |  | 3 |
| COL4A5 | Alport syndrome (COL4A5-related, X-linked) |  |  |  |  | 2 | 2 |
| CFTR | Cystic fibrosis | x | x | 2 |  |  | 2 |
| HBB | Beta-globin-related hemoglobinopathies |  | x |  | 1 |  | 1 |
| USH2A | Usher syndrome, type IIA |  | x |  | 1 |  | 1 |
| CYP21A2 | Congenital Adrenal Hyperplasia due to 21-Hydroxylase Deficiency |  | x |  | 1 |  | 1 |
| ATRX | Alpha-thalassemia mental retardation syndrome |  |  |  |  | 1 | 1 |
| DHCR7 | Smith-Lemli-Opitz syndrome |  | x |  | 1 |  | 1 |
| HEXA | Tay-Sachs disease |  | x |  | 1 |  | 1 |
| EYS | Retinitis pigmentosa 25 |  |  |  |  | 1 | 1 |
| NPHS2 | Nephrotic syndrome (NPHS2-related) / steroid-resistant nephrotic syndrome |  |  |  |  | 1 | 1 |
| F9 | Factor IX deficiency (X-linked) |  | x |  | 1 |  | 1 |
| SMN1 | Spinal muscular atrophy | x | x | 1 |  |  | 1 |
| FAH | Tyrosinemia, type I |  | x |  | 1 |  | 1 |
| ATP7A,<br>ATRX | Menkes disease, alpha-thalassemia mental retardation syndrome |  |  |  |  | 1 | 1 |
| FMR1 | Fragile X syndrome |  | x |  | 1 |  | 1 |
| GBA | Gaucher disease |  | x |  | 1 |  | 1 |
| **Total** |  |  |  | **3** | **18** | **10** | **31** |

### Post-conception administration of CS-283 results in a higher rate of prenatal diagnosis of affected fetuses compared to SPEP results

We obtained data for 279 ARCCs identified from a subset of MFM practices (out of the 4752 ARCCs we identified across all REI, MFM, and Ob-Gyn clinics in our study), and from these data we identified 246 (88%) who underwent fetal diagnostic testing as a consequence of the ARCC status resulting from administration of the CS-283 carrier screening test; the remaining 33 cases had canceled or incomplete fetal diagnostic testing results and so were excluded from further consideration. Of the 246 fetal diagnostic tests conducted, 24% (N=59) had a monogenic disorder genotype that was consistent with the disorder for which the ARCC had been identified as being at risk. Of the 59 positive cases, 69% (41 out of 59) corresponded to genes represented on the ACMG gene panel, while 24% (14 out of 59) corresponded to genes specific to the CS-283 gene panel (not in SPEP or ACMG), compared with only 4 cases that would have been identified by SPEP. Thus, the pan-ethnic CS-283 panel achieved a detection rate that was more than 13-times higher than that achieved with SPEP (Table 3).

**Table 3:** Fetus with confirmed diseases from at-risk carrier couples

| Genes | Disease Name | SPEP | ACMG | #_SPEP | #_ACMG | #_CS283 | Total |
| --- | --- | --- | --- | --- | --- | --- | --- |
| HBB | beta-globin-related hemoglobinopathies |  | x |  | 10 |  | 10 |
| GBA | Gaucher disease |  | x |  | 5 |  | 5 |
| CFTR | cystic fibrosis | x | x | 4 |  |  | 4 |
| ACSF3 | combined malonic and methylmalonic aciduria |  |  |  |  | 3 | 3 |
| GJB2 | non-syndromic hearing loss (GJB2-related) |  | x |  | 3 |  | 3 |
| USH2A | Usher syndrome, type IIA |  | x |  | 2 |  | 2 |
| MEFV | familial Mediterranean fever |  |  |  |  | 2 | 2 |
| ACADM | medium chain acyl-CoA dehydrogenase deficiency |  | x |  | 2 |  | 2 |
| CYP21A2 | congenital adrenal hyperplasia due to 21-hydroxylase deficiency |  | x |  | 2 |  | 2 |
| F11 | factor XI deficiency |  |  |  |  | 2 | 2 |
| HEXA | Tay-Sachs disease |  | x |  | 2 |  | 2 |
| NPHS2 | nephrotic syndrome (NPHS2-related) / steroid-resistant nephrotic syndrome |  |  |  |  | 1 | 1 |
| MCCC2 | 3-methylcrotonyl-CoA carboxylase deficiency (MCCC2-related) |  | x |  | 1 |  | 1 |
| RAPSN | congenital myasthenic syndrome (RAPSN-related) |  |  |  |  | 1 | 1 |
| CPT2 | carnitine palmitoyltransferase II deficiency |  | x |  | 1 |  | 1 |
| NEB | nemaline myopathy 2 |  | x |  | 1 |  | 1 |
| F9 | factor IX deficiency (X-linked) |  | x |  | 1 |  | 1 |
| PAH | phenylalanine hydroxylase deficiency |  | x |  | 1 |  | 1 |
| BBS12 | Bardet-Biedl syndrome (BBS12-related) |  |  |  |  | 1 | 1 |
| CYP27A1 | cerebrotendinous xanthomatosis |  | x |  | 1 |  | 1 |
| CFTR,DMD | cystic fibrosis; Duchenne muscular dystrophy / Becker muscular dystrophy (X-linked) |  | x |  | 1 |  | 1 |
| CYP21A2,MTM1 | congenital adrenal hyperplasia due to 21-hydroxylase deficiency; myotubular myopathy 1 (X-linked) |  |  |  |  | 1 | 1 |
| CYP21A2,BCKDHB | congenital adrenal hyperplasia due to 21-hydroxylase deficiency; maple syrup urine disease, type 1b |  | x |  | 1 |  | 1 |
| NPHS1 | nephrotic syndrome (NPHS1-related) / congenital Finnish nephrosis |  | x |  | 1 |  | 1 |
| CEP290 | Leber congenital amaurosis 10 and other CEP290-related ciliopathies |  | x |  | 1 |  | 1 |
| OTC | ornithine transcarbamylase deficiency (X-linked) |  | x |  | 1 |  | 1 |
| ATP7B | Wilson disease |  | x |  | 1 |  | 1 |
| PKHD1 | polycystic kidney disease, autosomal recessive |  | x |  | 1 |  | 1 |
| HPS1 | Hermansky-Pudlak syndrome, type 1 |  | x |  | 1 |  | 1 |
| TPP1 | neuronal ceroid-lipofuscinosis (TPP1-related) |  |  |  |  | 1 | 1 |
| IDS | mucopolysaccharidosis type II (X-linked) |  |  |  |  | 1 | 1 |
| IDUA | mucopolysaccharidosis type I |  | x |  | 1 |  | 1 |
| GLB1 | mucopolysaccharidosis type IVb / GM1 gangliosidosis |  |  |  |  | 1 | 1 |
| **Total** |  |  |  | **4** | **41** | **14** | **59** |

### Annualized economic burden of diseases represented in the CS-283 panel

Complementing impacts on reproductive health choices and pregnancy outcomes are potential healthcare savings through the prevention of pregnancies resulting in infants with severe monogenic disease. Diseases related to genes on the CS-283 panel can often lead to severe conditions that require extended treatment and costly care. To explore these costs and the impact comprehensive carrier screening could have on reducing overall healthcare burden and treatment costs, we built disease-specific cost models to estimate the economic burden imposed by the CS-283 diseases, and then weighted these cost models by inheritance patterns and individual carrier and ARCC rates specific to each of the 283 diseases. To capture the expected costs spread out over many diseases as a function of individual carrier and at-risk carrier couple rates, we computed the expected disease cost for pseudo-panels of size N that contained the N most common genes represented in the ARCCs, with N ranging from 1 to 283. To construct the pseudo-panels, we sorted the genes on the CS-283 panel by the empirical ARCC rates derived from our study (Supplementary Table 7). In cases where a gene did not give rise to a sufficient number of ARCCs to estimate a rate in our study, we estimated a theoretical ARCC rate based on random coupling across the 7 major population groups considered in our study. For each pseudo-panel, we estimated the overall ARCC rate for the panel (Figure 5A, red line, inner-most right-hand axis) and calculated the individual carrier rates (Figure 5A, blue line, left hand axis). The individual carrier rates rise more slowly than the ARCC rate due to X-linked genes such as *FMR1*, which are relatively more prevalent in ARCCs than individual carriers.

**Figure 5:**
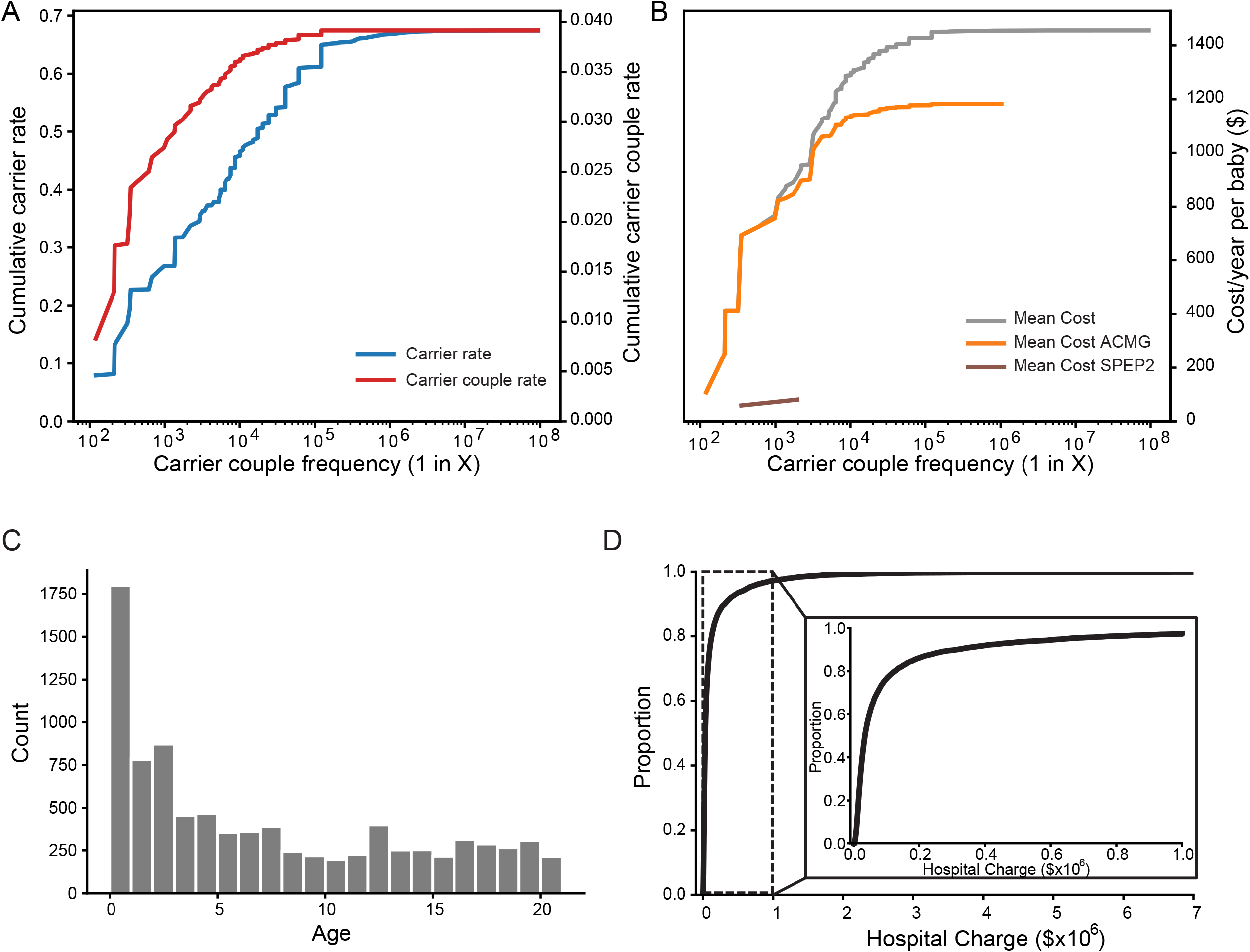
Economic burden of disease. **(A)** The genes in the CS-283 panel are ordered by ARCC frequency to estimate cumulative carrier rate (blue), at-risk carrier couple rate (red). **(B)** Single year disease-related in-patient hospital charges, based on the mean of the charges. The curves are generated incrementally for pseudo-panels with the N most frequent diseases, N ranging from 1 to 283 as described in the text. Note that there is an overlap between the CS-283 mean cost (grey line) and ACMG mean cost (orange line) initially due to intersection among genes in the two panels. **(C)** Distribution of age of patients in the CS-283 disease specific KID data cohort. **(D)** Cumulative distribution of individual hospital charges for patients in the CS-283 disease specific KID data cohort. *Inset*, Zoomed in cumulative distribution for charges less than $1m.

To estimate disease-specific costs, we constructed cost models for each disease using the 2019 version of the Kids’ Inpatient Database (KID), developed by the Healthcare Cost and Utilization Project (HCUP)^18^. This dataset contains hospital charge and diagnosis information (based on ICD-10 codes) for inpatient visits in the year 2019 for patients from 0-20 years old for all diagnoses. Therefore, the data provide an estimate of annual disease-related costs across all children. To estimate the cost for each disease we first mapped each disease caused by a gene in the CS-283 panel to an ICD-10 code (Supplementary Table 8). We found that all 283 genes represented on the CS-283 panel could be mapped unambiguously to an ICD-10 code for the corresponding monogenic disease. We then selected a cohort of patients from the KID Database whose primary diagnosis was in this list of 163 diseases (Supplementary Table 8). The resulting cohort had 8,882 patients (53.8% male), with a mean age of 6.8 years and a median age of 5.0 years (Figure 5B). The mean hospital charge across all CS-283 diseases was $139,805 (Figure 5C). We report the mean given some severe conditions require substantial resources, resulting in long tails in the distribution of hospital charges (Figure 5D). Therefore, the mean may provide a better picture of the overall cost of disease.

With the inheritance pattern, carrier and carrier couple rates, and cost estimated for each disease represented in the CS-283 panel, we combined these components to compute the expected cost for each pseudo-panel (See Methods). Our disease-specific cost model estimates the expected annual economic burden of the disease for any child, without requiring knowledge of the disease status. The expected cost for each pseudo-panel is the sum of the disease-specific costs for each gene in the pseudo-panel (Figure 5A, gray line for the mean cost). The mean cost rises more slowly than the ARCC rate for the more prevalent genes, then more rapidly in the middle of the curve (between ARCC frequency of 1 in 1,000 to 1 in 10,000). This suggests that the economic burden of the diseases with ARCC frequencies between 1 in 1,000 and 1 in 10,000 may be underestimated when considering ARCC rates alone. Our model estimates the potential economic burden associated with those pediatric genetic diseases covered by the CS-283 panel. In total, the 2019 KID database identified 8,882 patients who were hospitalized with a primary ICD-10 diagnosis matching the CS-283 definition of pediatric genetic disease. When averaged across a population of newborns (healthy and affected), our model estimates that the annual direct medical costs identified by the diseases on the full CS-283 panel is $1,477 per child. In comparison, diseases identified by the ACMG panel detect $1,206 per child. However, in sharp contrast, the two diseases represented on the SPEP only captures 5.5% ($81) of the annual cost of disease compared to the CS-283 panel ($1,477); and 6.7% of costs compared to the ACMG panel ($1,206).

## Discussion

Despite increasing scales of genomic data generated in the research and clinical settings that have driven dramatic increases in our understanding of monogenic and complex forms of disease, and despite the increasing demonstrations of clinical utility comprehensive genomic profiling data exhibits across a broad spectrum of diseases and conditions, in the reproductive health setting the routine screening of patients to determine carrier status for severe monogenic disorders to enhance reproductive health choices, has not achieved widespread clinical adoption and coverage by health insurers. To add to a growing body of evidence demonstrating the clinical utility of pan-ethnic comprehensive carrier screening, here we provide one of the largest studies published to date examining more than 390,000 patients who have received a 283-gene, comprehensive carrier screening test, resulting in the identification of roughly 270,000 individual carriers and more than 4,000 at-risk carrier couples (ARCCs). With nearly ten times the number of ARCCs identified in our study compared to other published studies, we were able to examine outcomes in a subset of the ARCCs with respect to embryo selection and implantation, and pregnancy outcomes.

Our study is differentiated in several ways from previously published expanded carrier screening studies with respect to the levels of evidence we provide that support comprehensive pan-ethnic carrier screening for all pregnancies. First, our study provides results from one of the most comprehensive carrier screening panels comprised of 283 genes, now run on 397,540 patients, resulting not only in carrier detection rates > 60% across all major population groups, but in achieving one of the largest ARCC rates reported to date. When these results are compared to the current standard pan-ethnic carrier screening panel comprised of 2 genes that cover the higher frequency, severe conditions cystic fibrosis and spinal muscular atrophy, greater than 12-times the number of ARCCs are identified with CS-283 compared to SPEP. Second, for more than 142,049 of the patients tested on the CS-283 panel, low pass whole genome data were generated to infer genetic ancestry, resulting in one of the largest test populations with genetic and self-reported ancestries in the context of comprehensive carrier screening. Recent ACMG guidelines recommend carrier screening for all pregnancies across 113 genes with a carrier frequency of 1 in 200 or higher and X-linked conditions, but they also highlighted that the recommended list of genes may not adequately address variations in allelic frequencies associated with ancestry^23^. Low pass WGS provides a potential solution to infer genetic ancestry in patients undergoing carrier screening without the need to rely on self-reported ancestries. In addition, lpWGS has several distinct advantages compared to comprehensive genotyping microarrays with respect to more accurate identification of genetic variation across the allele frequency spectrum, particularly in underrepresented populations^39^. From a pragmatic point of view, our use of lpWGS data to decipher the complex structure of genetic ancestry in our test population highlights the significant discordance between self-reported and genetic ancestries, raising questions as to whether self-reported ancestries are reliable enough to forego more comprehensive carrier screening panels that can substantially impact test results and ultimately pregnancy outcomes, and if coverage of more comprehensive panels should be inclusive on the basis of self-reported ancestries alone. For example, insurance coverage for carrier screening beyond a minimal SPEP panel often depends on meeting ancestry requirements, such as having ASJ ancestry derived from a second or first degree relative of ASJ descent^27^. However, of the 12,612 patients we determined had genetic ASJ ancestry to the degree required by major healthcare insurers to warrant coverage for carrier screening using an ASJ-specific screening panel^27,40^, only 2,409 self-reported with ASJ ancestry, leaving 10,203 patients (80.9% of the ASJ population in our test population) who would otherwise qualify for enhanced carrier screening, receiving a known substandard of care with SPEP.

Third, given the significantly enhanced benefit with respect to identification of carriers and at-risk carrier couples, the comprehensive pan-ethnic carrier screening panels (ACMG and CS-283) provide to specific population groups over ancestry-specific panels designed specifically for a given population group, our results directly support the ACMG’s recommendation that “Carrier screening paradigms should be ethnic and population neutral and more inclusive of diverse populations to promote equity and inclusion.”^23^ In fact, this result is consistent with the evolution of screening for cystic fibrosis from ethnicity-specific screening to population-wide screening. While cystic fibrosis is most common in non-Hispanic Caucasian populations, with a disease incidence of 1 in 2,500 and a carrier rate of 1 in 25 ^41^, in 2005 ACOG changed its initial recommendation of offering CF carrier screening to high-risk ethnicities only, to making CF the standard of care for all patients regardless of ethnicity, given the increasing difficulty of assigning a single ethnicity to any given individual ^42^.

A fourth differentiator of our study is the linkage of a subset of the ARCCs identified in our test population to REI and pregnancy outcomes that were directly impacted by the CS-283 carrier screening results, significantly beyond what would have been achieved with a standard pan-ethnic or ancestry-specific carrier screening panels. In the REI setting, positive carrier screening results involving genes that were not part of the SPEP, motivated preimplantation genetic testing of embryos generated from the corresponding ARCCs, with a substantial percentage of the embryos testing as positive for severe monogenic disease genotypes. As a result, rather than disease-causing embryos getting implanted and leading to pregnancies that result in an infant with a severe monogenic condition, the disease embryos were not selected for implantation. Similarly, in the MFM setting, a subset of ARCCs identified after becoming pregnant, had prenatal diagnostic testing carried out on the fetus, which in a significant percentage of the cases (roughly 25%) led to a positive diagnosis of the disease genotype in the fetus, thus expanding the reproductive choices that could be considered in the pregnancy moving forward. In a majority of these cases in which embryos or fetuses were diagnosed as having a severe monogenic disease genotype, such diagnoses would not likely have been made if an SPEP carrier screening test had been employed, given our results depended on the broader set of disease conditions represented on the CS-283 panel.

A final differentiating feature of our study was the modeling of the economic burden of diseases covered by the CS-283 panel as well as all sub-panels of ranging in size from one gene to 283 genes, with each panel of size N in this range comprised of the N genes with the highest ARCC rates. Our modeling incorporated empirically estimated carrier and at-risk carrier couple rates from our test population, in addition to empirical cost of care estimates across all the disease conditions represented in the AHRQ database and overlapping with the CS-283 panel. The results from our model demonstrated the potential for significant savings directly supported by our outcomes data that could be realized through the delivery of comprehensive pan-ethnic carrier screening as the standard of care for all. Such projected savings would result from improved pre-pregnancy planning enabled by a couples’ knowledge of their risk for delivering offspring with a severe monogenic disorder. Our cost model estimated a conservative economic burden of $1,477/year for diseases represented on the CS-283 panel for every child in the U.S. regardless of disease status. That is, the cost of treating children with severe monogenic disorders represented on the CS-283 panel, when spread out across all children in the U.S., is $1,477 per year per child. However, SPEP only captures 5.5% ($81) of the annual cost of disease compared to the CS-283 panel. Therefore, the full CS-283 panel provides more opportunities to (1) reduce the diagnostic odyssey; (2) avoid costs associated with unnecessary testing; and (3) treat disease before advanced progression. Reducing pregnancies that result in the delivery of infants with these severe monogenic disorders thus provides for the potential to save many billions of dollars per year in health care costs. We note that with our modeling, the economic burden of pediatric genetic diseases may be underestimated when considering ARCC frequencies less than 1-in-200. For example, a recent study that employed a phenotypic approach to characterize US pediatric patients with clinical indicators of genetic diseases relied on >2,000 ICD-10 codes to identify cases from commercial payer data with possible, probable, or definite genetic causation^43^.

From a genomic perspective, our results support previous findings that the marginal cost of screening and detecting carriers does not exceed the marginal benefit of detecting at-risk couples^44^. Further, the clinical benefits associated with identification of carriers and testing of their partners outweigh the costs of including rare conditions on a comprehensive carrier screening panel. Our results suggest that the 1-in-200 carrier rate threshold will limit detection of at-risk couples associated with significant downstream clinical and economic burden (as supported in Figure 5A). While cost is not explicitly considered as a measure of severity, it should be noted that charges derived from annotated hospital billing data reflect healthcare resource use intensity, particularly in the NICU, where charges capture mechanical ventilation, surgeries, and insertion of feeding devices. Costs should therefore be considered as a surrogate for severity. Our data and modeling support that the severity of a rare monogenic disorder should be considered as a criterion for inclusion on carrier screening panels in addition to carrier frequency.

Despite our best efforts, several limitations exist in our study. A large portion of our study cohort (60%) did not have self-reported race/ethnicities, given patients are not required to provide this personal information when their physician orders the tests. Such under reporting hindered the precision for ordering the most appropriate carrier screening test based on the patient’s ethnicity, and in addition created challenges for our study. Among the patients with self-reported ethnicities, over 99% of the patients reported only a single ancestry group, which may not reflect the ancestry admixture that is relatively common in the US population. We attempted to reduce the impact of this under reporting by utilizing lpWGS data along with CS-283 testing to infer genetic based ancestries for patients. Even though we only had roughly 35% of the individuals in our test population with genetic ancestry data, our findings can be considered robust given the relatively large sample size achieved across hundreds of REI and MFM centers. Another limitation is that the CS-283 panel was designed nearly 5 years ago, and thus has expected deficiencies in coverage given the pace of genomic information growth. For example, CS-283 covers only 77 of the 113 “tier 3” genes recommended by ACMG guidelines as a standard of care for all^23^. However, we note that CS-283 ranks among the top 3 of the 7 commercially available comprehensive carrier screening panels available today in terms of coverage of the ACMG 113 gene set (Supplementary Figure 1).

With respect to the outcomes analyses, we described the sensitivity and specificity of using International Classification of Disease 1^0t^h Edition (ICD-10) codes for preimplantation or prenatal genetic diagnosis (PGD), something that has been previously described in real-world research ^17,18^. Some PGDs were matched to the closest ICD-10 code, which may capture a broader group of people than we intended. Conversely, highly sensitive ICD-10 codes may exclude some patients with disease. For pediatric genetic diseases, the dearth of longitudinal cohorts with sufficient sample sizes makes it difficult to estimate disease-specific penetrance and incidence rates^17^. While double counting may exist (e.g., an individual patient with multiple, separate admissions), the database curation methods have minimized this at scale for newborns leading to NICU stays and ER admissions leading to hospitalizations. Finally, the hemoglobinopathies represent a group of conditions characterized by abnormalities of alpha or beta chain production or oxygen affinity. The use of molecular testing is associated with a greater relative risk compared with hemoglobin electrophoresis in screening for hemoglobinopathies, because the pathogenic variants that result in β-thalassemia are often private (specific to families). Furthermore, the genomic changes associated with α-thalassemia are deletions and the most severe α-thalassemia phenotype requires deletions in cis (on the same chromosome). Although NGS can determine dosage (the number of copies of a gene), a limitation of current short-read NGS technologies is the inability to determine cis or trans (on opposite chromosomes)^45^.

In conclusion, this study provides further evidence of the utility of pan-ethnic CS as recommended in the updated ACMG guidelines by informing reproductive partners about a much wider array of reproductive health risks. The CS-283 gene panel identifies significantly more carriers and carrier couples compared to ancestry-based screening, most notably in non-Caucasian populations, such as SAS, EAS, AFR, and AMR; population groups that received the greatest benefit. As a result, we can conclude from our results that comprehensive, pan-ethnic CS over traditional ethnicity-based screening leads to improved health equity in these relatively understudied populations. Future studies should evaluate both the provider and patient experience with comprehensive pan-ethnic carrier screening in the pre-pregnancy and prenatal periods. Additional studies are needed to explore the patient’s experience in learning genetic ancestry via CS in conjunction with lpWGS to aid in patient involvement of these advancing technologies.

## Data Availability

All original carrier screening results data are from Sema4 genetic counselor reports. Carrier rate and ARCC rate summary statistics are available in Supplementary Table 5 and Supplementary Table 7.

## Acknowledgement

We thank Jay Shaw (genetic counselor) for interpreting the CS reports. We also thank Sema4 IT and health informatics staff, Shay Hassidim, Pandian Pandian, and Alvin Chen, for supporting raw CS data delivery.

## Author contribution

L.L, R.C, and E.S designed the study. R.A.S, M.K.H, Y.Z, J.T, L.Y.L, S.L participated in the data analyses. R.C supported the data delivery for the analysis. T.A.C and A.B.C provided embryo data from Reproductive Medicine Associates of New York. A.B, B.W, M.F, L.S and L.E provided genetic insights for CS from the lab. All authors participated in the data analysis, data interpretation, manuscript writing, and approval of the final submitted version.

## Data Source/Availability

All original CS results data are from Sema4’s genetic counselor reports. Carrier rate and ARCC rate summary statistics are available in Supplementary Table 5 and Supplementary Table 7.

## Figure Legends

**Supplementary Figure 1:** Gene coverage across 7 available comprehensive CS tests and ACMG guideline. 1A) Sema4 CS-283 vs others. 1B) Sema4 CS-502 vs others.

Overlapped genes: Sema4-283: 77 genes (27.2% coverage of ACMG), Sema4-502: 86 genes (17.1% coverage of ACMG), Natera-274: 73 genes (26.6% coverage of ACMG), Invitae-302: 79 genes (26.1% coverage of ACMG), Fulgent-335: 80 genes (23.9% coverage of ACMG), LabCorp-143: 52 genes (36.4% coverage of ACMG), Myriad-176: 66 genes (37.5% coverage of ACMG), GeneAware-159: 60 genes (37.8% coverage of ACMG). Source: https://www.ncbi.nlm.nih.gov/gtr/

## Materials and Methods

### Study Population

Our cohort consisted of 397,540 (236,355 females) patients who were tested from January 2019 to July 2022 using next generation sequencing, expanded carrier screening panels with a common core set of 283 genes. The expanded carrier screening panels were developed and run by Sema4 (http://www.sema4.com) on all patients. Patients were included in this study only if they were tested using an NGS panel containing the core set of 283 genes (CS-283). For patients who received screening on larger panels, only the results for genes in the CS-283 panel were included in this analysis. Patient information was de-identified prior to all analyses.

In addition to the carrier screening results, we also collected information on the practice where the test was ordered and classified each practice as either a Reproductive Endocrinology and Infertility practice providing in vitro fertilization (REI), a Maternal Fetal Medicine (MFM) practice, or a general Obstetrician-gynecologist practice (Ob-Gyn). Of the 397,540 patients, 306,226 ordered tests through 332 REI practices, while the remaining 91,314 were from 745 MFM/Ob-Gyn practices. In addition, we mapped each practice’s location in the U.S. to the county level to show the geographic diversity of our test population.

Patients’ self-reported ethnicity was captured in a free text field on the test order form. When this field was left blank, the patient was assigned “Not reported” (NR) ethnicity. This field was not required for the carrier screening test to be ordered. Thus, an NR ethnicity designation may be due to an omission by the ordering physician even when the patient ancestry is known, uncertainty by the ordering physician on the patient’s ancestry, the patient not knowing their ancestry, or the patient deciding not to report their ancestry. To avoid any potential confounding factors due to the possible reasons for having not reported ethnicity, we excluded these NR patients when performing self-reported ethnicity-based analyses.

Finally, from the patient medical data we assembled, we had information about patient relationships and thus were able to link the carrier screening results of reproductive partners when this information was present in the medical record. We were able to identify the male reproductive partner for 121,882 out of the 236,355 females screened (51.6%) in or test population. Couples who were carriers for the same autosomal recessive condition or where the female was a carrier of an X-linked condition were considered At-Risk Carrier Couples (ARCC) (Figure 1A, 1E, Supplementary Table 1).

### Carrier screening results extraction

Results from carrier screening were extracted from notes written by genetic counselors (GC), which included interpretation of the test results. We applied rule-based natural language processing (NLP) on the GC notes to identify gene, genetic variants, and zygosity. Only variants considered to be pathogenic or likely pathogenic were reported on Sema4’s carrier screening panels. A healthy patient is considered to be a carrier if they were identified to be heterozygous for one pathogenic/likely pathogenic variant in an autosomal recessive disease gene or if either a healthy female or male is hemizygous for a pathogenic/likely pathogenic variant in an X-linked disease gene.

For some genes on the CS-283 panel, combinations of pathogenic variants found in trans (for distinct variants) or homozygous in patients are unlikely to cause disease with that specific combination of variants (“silent carriers”; Supplementary Table 2), although in combination with other pathogenic variants could cause disease. This information was taken into account for identifying ARCCs. That is, if the combination of pathogenic variants in a carrier couple that could be passed on to offspring would lead to a “silent carrier”, then we did not consider the carrier couple as an ARCC. We also note that 20 variants in *CYP21A2* are considered mild, giving rise to non-classic congenital adrenal hyperplasia (NC-CAH) when two mild alleles occur in *trans* or one mild allele and one classic allele occur in *trans* (Supplementary Table 3)^46^. Carrier couples with *CYP21A2* variants that would lead to NC-CAH, but not to more severe forms of CAH, were included for the overall ARCC counts.

Additionally, alpha thalassemia is unique in that four α-globin genes are present that may be deleted or silenced leading to various phenotypes^47^. For patients who were identified as carriers of *HBA1/2* in the notes, we extracted the copy numbers. Individuals with a genotype of αα/α-in *HBA1/2* are not at risk of having offspring with an alpha thalassemia-related disease and thus were considered as α- thalassemia silent carriers^47^. At-risk carrier couples (ARCC) were also determined by copy numbers present in each partner. We considered the following conditions as ARCC for alpha thalassemia: 1. A α-thalassemia trait carrier in *cis* (αα/−−) and an α-thalassemia trait carrier in *trans* (α−/α-), leading to a 50% risk of developing Hemoglobin H (HbH) disease (−−/−α); 2. Two full carriers in *cis* (αα/−−), or a patient with HbH (α−/−−) and an α-thalassemia trait carrier in *cis* (αα/−−), or both patients with HbH (α−/−−), leading to a 25% risk of developing Hemoglobin Bart hydrops fetalis (−−/−−)^47^. It’s known for certain very rare combinations of point mutations in *HBA1* or *HBA2* and deletion of *HBA1/HBA2* can cause severe disease. For this reason, such combinations we identified in the couples (11 in total) have been manually reviewed by Sema4 genetic counselors. All 11 combinations present in our analysis cohort have been concluded to be negative to cause disease.

### Definition of three screening panels

We compared both individual carrier rates and ARCC rates across 3 carrier screening panels motivated by professional guidelines and medical policies from health insurers. The first panel was comprised of 2 genes that are generally supported by ACOG guidelines and by health insurers’ medical policies for use in all pregnancies. Two genes were reaffirmed by ACOG in 2020 as part of pan-ethnic screening in all patients (*CFTR and SMN1)*^*35*^. We refer to this 2-gene panel as the standard pan-ethnic panel (SPEP). The second panel is comprised of 77 genes (*ABCC8, ABCD1, ACADM, ACADVL, ACAT1, AGA, AGXT, AIRE, ALDOB,ALPL, ARSA,ASL,ASPA,ATP7B,BBS1, BBS2, BCKDHB, BLM, BTD, CBS, CEP290, CHRNE, CLRN1, CNGB3, COL7A1, CPT2, CTFR, CYP21A2, CYP27A1, DHCR7,DHDDS, DLD, DMD, ELP1,F9, FAH, FANCC, FKRP, FKTN, FMR1, G6PC, GAA, GALT, GBA, GBE1, GJB2, GLA, GNPTAB, HBA1/2, HBB, HEXA, HPS1, HPS3, IDUA, MCCC2, MCOLN1, MLC1, MMACHC, MMUT, NEB, NPHS1, OTC, PAH, PCDH15, PKHD1, PMM2,RARS2,RS1,SLC26A2, SLC26A4, SLC37A4, SLC6A8, SMN1, SMPD1, TMEM216, USH2A*) that are recommended by ACMG guidelines and included in our CS-283 panel^48^. The third panel represents a set of 283 genes included on Sema4’s most comprehensive expanded carrier screening products and is referred to as CS-283. The panel is comprised of genes associated with diseases in which the carrier frequency is more common than 1 in 500 in the general population or in a subpopulation (e.g., Ashkenazi Jewish), and where the disease characteristics meet: 1) early-onset and severe; 2) childhood/early adult onset and progressive, or 3) amenable to treatment or intervention. The genes included in CS-283 panel are given in Supplementary Table 4.

### Genetic ancestry specific panel construction

Carrier frequencies (Cf) for the genes included in this study have been curated by Sema4 internally, mainly utilizing The Human Gene Mutation Database^49^ and Gnomad^50^. Supplementary Table 9 has full information on Cf.

To construct the ancestry specific gene panel, Supplementary Table 9 was used. We used 1 in 100 Cf as cutoff and applied to each ancestry specific tab. Supplementary Table 9 is sorted by Cf from largest to smallest. Any genes with Cf >= 0.01 have been included as the ethnicity specific/genetic ancestry specific gene panel.

### Real-world evidence from REI and high-risk population in obstetric clinics

To validate whether pathogenetic genetic variants translate from ARCC to their embryos or fetus and assess the impact of ARCC status on reproductive decision-making, we obtained two real world evidence data sets for investigation that included subsets of patients from our full test population. The first set was comprised of 117 ARCCs identified at the Reproductive Medicine Associates of New York (RMA-NY), a Manhattan-based fertility clinic. Of these 117 ARCCs, 43 underwent Preimplantation Genetic Testing for Monogenic Disorders (PGT-M) from Jan 2019 to March 2022. We obtained the embryo and patient outcome data for evaluation. The second set was comprised of 279 ARCCs identified in a subset of MFM/Ob-Gyn practices in which patients were tested using the CS-283 panel and pursued prenatal diagnostic test from Jan 2019 to March 2021. Of the 279 ARCCs, 88% (246/279) underwent chorionic villus sampling and/or amniocentesis and had fetal diagnostic testing conducted, and we retrieved the results of this testing for further assessment. The remaining 12% of ARCCs had incomplete or canceled diagnostic testing and so were excluded from further analyses. We retrieved fetal diagnosis data for further assessment.

### Cost model

We utilized the Kids’ Inpatient Database (KID) which lists inpatient costs in 2019, developed by the Healthcare Cost and Utilization Project (HCUP), sponsored by the Agency for Healthcare Research and Quality (AHRQ), referencing previous methods developed with the 2012 version of KID.^18^ The KID Inpatient Database is based on administrative data such as discharge abstracts created by hospitals for billing. The KID is the largest publicly available all-payer pediatric inpatient care database in the United States, containing approximately 3 million pediatric discharges each year from 48 HCUP member states plus DC. KID data is sampled from ~4,000 U.S. community hospitals (defined as short-term, non-Federal, general and specialty hospitals, excluding hospital units of other institutions) and excludes rehabilitation hospitals.

Since 1997, AHRQ has released KID datasets in three-year cycles (1997, 2000, 2003, 2006, 2009, 2012, 2016). Several studies have analyzed KID datasets for newborn and pediatric healthcare resource utilization, ranging from rare diseases to highly prevalent chronic conditions^51–55^. In the 2019 KID dataset, 51.4% are female. Self-reported race is Caucasian (45.6%), African American (16.6%), Hispanic (19.7%), Asian/Pacific Islander (4.0%), and Native American (0.9%). The primary payers are Medicaid (50.8%), private insurance (41.2%), and self-pay (4.3%). Small counties with populations <250,000 comprise 21.9% of the patient sample.

Healthy newborns receiving routine care were sampled at a rate of 10%, while newborns with complicated courses and other pediatric discharges (age 20 or less at admission) were sampled at a rate of 80%. Pediatric discharges cover all payers, including children covered by Medicaid, private insurance, and the uninsured. The large sample size enables analyses of rare conditions and uncommon treatments.^18^ Among cases with genetic diseases matched to corresponding ICD-10 diagnostic codes, 49.6% are female. Self-reported race is Caucasian (45.9%), African American (20.6%), Hispanic (21.5%), Asian/Pacific Islander (4.0%), and Native American (1.1%). The primary payers are Medicaid (53.3%), private insurance (38.6%), and self-pay (2.9%). Small counties with populations <250,000 comprise 20.1% of the patient sample.

We estimated the expected hospital-related costs attributable to the diseases on the CS-283 panel for any new child born using the following equation

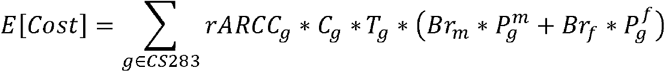

*E*[*Cost*] is the expected cost for any new child born, independent of disease status. *rARcc*_*g*_ is the ARCC rate for gene g in the CS-283 panel. This ARCC rate was determined empirically if we had at least one couple in our data that were carriers of the gene. If there were no couples where both partners were shared carriers for *g*, we assumed random coupling across ethnicities and estimated the ARCC rate as the square of the individual carrier rate for non-X-linked genes, and as the individual carrier rate for X-linked genes. *C*_*g*_ is the mean cost estimated from the AHRQ data set from all patients whose primary diagnosis includes the ICD code for the disease caused by g. An expert medical geneticist reviewed a list of ICD-10-CM codes (May 2022), which enabled identification of codes for genetic diseases related to genes in the CS-283 panel. *T*_*g*_ is the transmission probability of passing on a disease genotype to an offspring. For autosomal recessive diseases (the vast majority of diseases represented on the CS-283 panel) the transmission probabilities are 0.25. We multiply the cost function by the transmission probability to account for the likelihood of an ARCC giving birth to a child with a disease genotype for gene *g*. Finally, 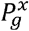 represents the average penetrance for gene *g* across all disease genotypes for individuals of sex *x* (either male or female) and is weighted by *Br*_*x*_, the probability of delivering a child of sex *x*. The penetrance for the diseases represented on CS-283 are generally close to 1.0. The cost function is multiplied by the weighted penetrance to account for the likelihood of a child manifesting the disease if they have the disease genotype.

### Low pass whole genome sequencing (lpWGS) raw sequencing data processing and base calling

Low pass whole genome sequencing was carried out as previously described^39^. First, we ran a quality control pipeline using raw paired end FASTQ files generated from the low pass whole genome sequencing runs. The minimum number of reads per individual was four million paired end read pairs, for a total of eight million reads. Second, we utilized bwa^56^ for the mapping step with reference genome version g1k_v37^15^. The minimum number of mapped reads was six million. Third, in the base calling step, we used *SAMtools*^*57*^ and the *mpileup* calling algorithm framework^58^. The output from this step is the input for the genetic ancestry inferences pipeline described below. The minimum number of mapped reads was six million. Third, in the base calling step, we used *SAMtools*^*57*^ and the *mpileup* calling algorithm framework^58^. The output from this step is the input for the genetic ancestry inferences pipeline described below. In total, we have 142,049 out of 397,540 individuals that have lpWGS based ancestry data in this analysis. We classify couples into a specific ancestry if either partner has a value of 25% or more of that ancestry component as determined by the lpWGS results; e.g. if a female partner has 0.35 ASJ, 0.65 NFE, and male partner has 0.9 NFE and 0.1 AFR, this couple will be classified to have both ASJ and NFE ancestry.

### Self-Genetic Ancestry Inferences

We used the output from processed lpWGS data (method above) run through Gencove’s proprietary ancestry inference workflow [https://www.gencove.com/products, https://docs.gencove.com/main/data-analysis-configurations/#human]. This ancestry inference workflow has been increasingly adopted as a more accurate method for genotyping and inferring ancestry^59,60^. Using this pipeline, we derived the following genetic ancestry population groups: African (AFR), Admixed American (AMR), Ashkenazi

Jewish (ASJ), East Asian (EAS), Finnish (FIN) European, Non-Finnish European (NFE), and South Asian (SAS). If a population group could not be confidently assigned or was ambiguous, an Unassigned/ambiguous category is assigned.

Any individual whose lpWGS-based ancestry proportion is greater than the threshold (25%) was classified as being of descent from the given ancestry category. In addition, we mapped self-reported ancestry to the genetic ancestry population groups. Self-reported ancestry was collected in a free text field on the test order form. The specificity of the individuals self-report varied from broadly geographic (e.g. “European”) to sub-country region (e.g. “Fujianese”). However, most reports were at the country level. In addition, most self-reports were in the form of a geographic location (e.g. “Brazil”) or the associated demonym (“Brazilian”). To map this self-reported information to the 7 broad population groups, we mapped each unique demonym to its associated country. Then we mapped each country to an ancestry population group based on geographical borders determined by Gencove (https://ancestry.gencove.com). If the country was split by a geographic border, we mapped to the ancestry population group comprising greater territory in that country. Finally, since the ASJ population comprises over 90% of the Jewish population in the United States^61^, we mapped any patient who identified as “Jewish” to the ASJ population group.

### Benefit-Gain analyses

To calculate the differences in detection rate when going from an ancestry-based panel to the CS-283 panel (or ACMG-informed panel), we used the following:

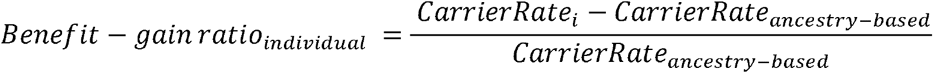

where *i* refers to one of the comprehensive pan-ethnic panels, CS-283 or the ACMG-informed panel (see Figure 4A). Analogously, we use the following to determine the ratio for at-risk carrier couples (see Figure 4B):

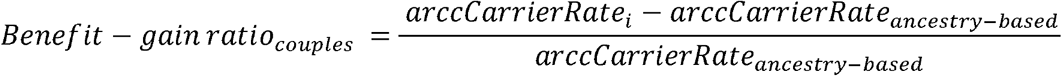

We received approval from Western Institutional Review Board (IRB#: 20215113) and Mount Sinai School of Medicine in accordance with Mount Sinai’s Federal Wide Assurances (FWA#00005656, FWA#00005651) to conduct this study.

