## Supplementary figures and images for "Real-world genetic screening with molecular ancestry supports comprehensive pan-ethnic carrier screening"

### Supplementary Figure 1

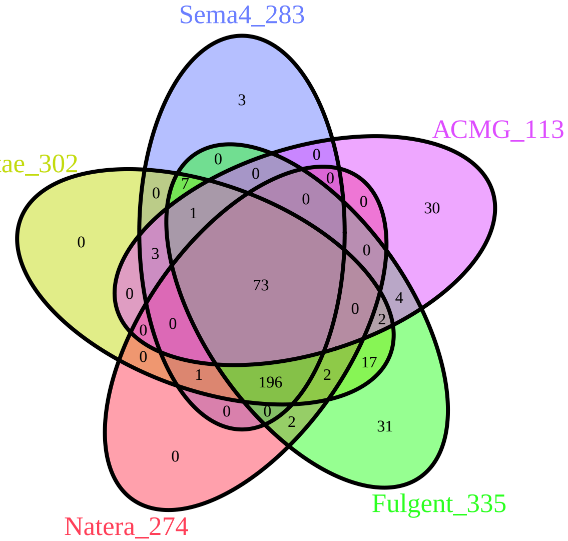

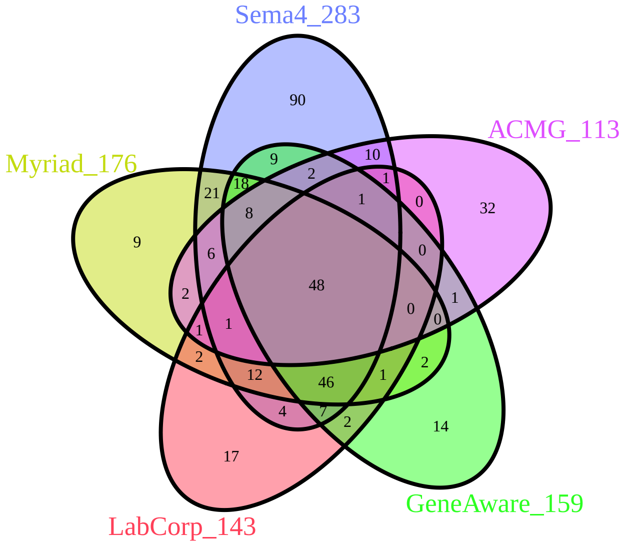

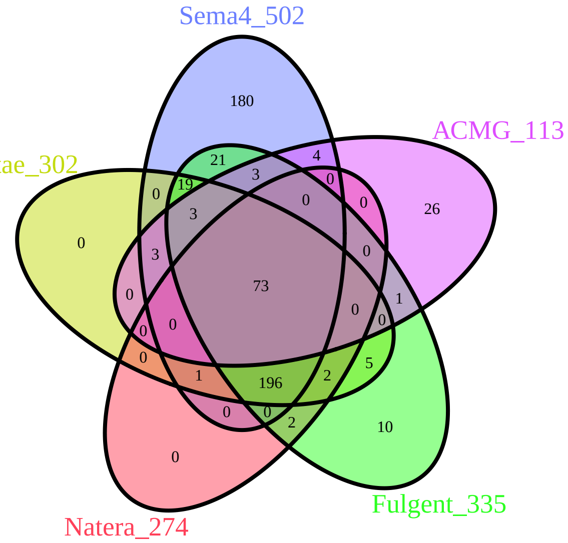

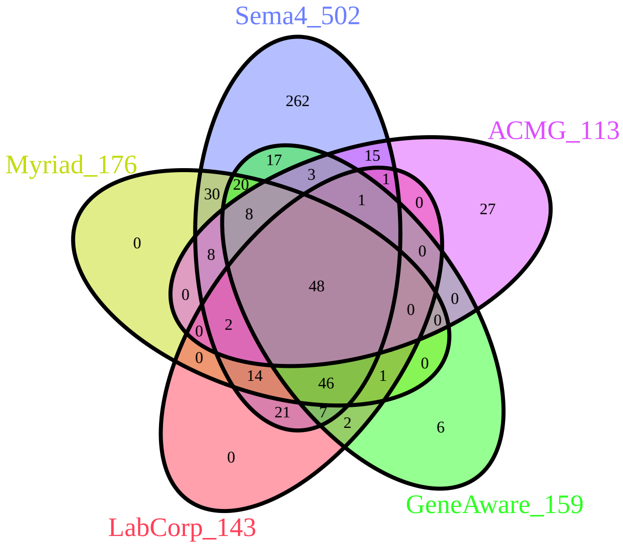
